# Study Protocol for Evaluation of The Impact of The Door-to-Door Approach by Community Health Workers and The Novel Mobile Application for Self-Reporting on HIV Testing Rates among Men-headed Households in Tanzania

**DOI:** 10.1101/2025.04.17.25326002

**Authors:** Hajirani M. Msuya, Kassimu Tani, Omar Lweno, Mwifadhi Mrisho, Bakar Fakih, Omar Juma, Ali Ali, Silas Temu, Gumi Abdallah, Hassan Tearish, Zeye Masunga, Werner Maokola, Neema Mpanduji, Moza Kassim, Ramadhani Abdul, Abdallah Mkopi

## Abstract

Sub-Saharan Africa faces a disproportionate burden of HIV/AIDS, and Tanzania, reflecting this trend, faces challenges with only 60.6% of individuals aged 15–64 aware of their HIV status. Nevertheless, according to the present state of affairs, the mean coverage rate of previous HIV testing in Tanzania (66.5%) is still behind the UNAIDS target. The UNAIDS 2030 target urges a 95% awareness rate among those infected, sustained ARV therapy, and viral suppression. To address this, Tanzania amended its HIV and AIDS Act, permitting voluntary HIV self-testing. The Ministry of Health (MoH) initiated the distribution of oral HIV self-test kits, involving community health workers (CHWs). The Ifakara Health Institute (IHI) and their partners will conduct the operational research program. The main objective is to evaluate the impact of the door-to-door distribution of oral self-testing kits by community health workers and the novel mobile application for self-reporting HIV testing results among men-headed households in Tanzania.

The program is set to be implemented in four regions namely; Tabora, Kigoma, Manyara, and Kaskazini Pemba in Tanzania. These regions had low prior HIV testing rates among men. CHWs will conduct door-to-door distribution of HIV self-test kits to male-headed households, emphasizing the use of the novel mobile application for self-reporting results. In this operational research program, we will use a cluster randomized controlled trial with regions’ wards as the clusters to evaluate a well-defined intervention using a door-to-door approach and a mobile App within intervention wards in rural and urban communities. The sample size calculation considers regions’ population sizes and prior HIV testing rates, aiming to achieve an increase in self-testing. Ethical considerations will emphasize confidentiality, written informed consent, and careful monitoring of potential adverse outcomes, focusing on gender-based violence prevention. The program timeline spans 24 months, encompassing preparation, training, pilot testing, baseline, and subsequent surveys, with an emphasis on data analysis, report writing, and dissemination in the final phase.

## Introduction

Sub-Saharan Africa continues to be the region most impacted by HIV/AIDS, which has remained a major global health concern where more than two-thirds of people living with HIV reside [1]. The current treatment target encouraged by UNAIDS in 2030 is to achieve 95% of infected individuals knowing their status, 95% of those diagnosed with sustained ARV therapy, and 95% of those receiving antiretroviral (ARV) therapy achieving viral suppression [2]. HIV testing is a crucial first step to accessing HIV prevention and treatment services [3]. According to the DHS-MIS Tanzania Report 2022, 80% of women between the ages of 15 and 49 have been tested for HIV and have been informed of the results of their most recent test. On the other hand, only 64% of men have had an HIV test and have received the results of their most recent test [4]. Early diagnosis offers a chance to provide persons living with HIV (PLHV) with knowledge and abilities to stop HIV from spreading [5].

A systematic review identified self-testing as one approach that could improve testing and counseling uptake for individuals who prefer to test with a degree of privacy [6]. One recently developed tool that may help us reach more individuals for HIV testing is the oral self-testing (HIVOST) kit. The oral self-testing for HIV has been used and approved in several African countries [7]; such as Kenya [8], Uganda [9], Malawi [10], Zambia [11], Zimbabwe [12], etc. This may provide an important opportunity to increase the proportion of individuals with HIV testing. First, the oral test does not require a finger prick, making self-administer easier. Second, the distribution of self-testing kits appears to break down numerous barriers to testing including the desire for privacy, fear of stigma around HIV, time required to be tested at a clinic, and control over the process [13]. However, the concern is that the individual who uses an HIV self-testing kit and tests positive must follow the necessary steps, including pre-test counseling, post-test counseling, and linkage to care.

Community health workers (CHWs) could distribute the HIVOST kits in the community [14]. The CHW workforce is an integral part of primary healthcare services. A lack of formal CHW engagement has prevented the achievement of universal healthcare goals [15]. Generally, CHWs perform different functions [16]. A systematic review indicated that CHWs can contribute to HIV services delivery and strengthen human resource capacity in sub-Saharan Africa [17].

In 2019, Tanzania launched a campaign for HIV Testing countrywide including HIV self-testing. The HIV and AIDS (Prevention and Control) Act, Cap. 431 has been amended to allow HIV Self-Testing (HIVST) [18]. The law requires HIV self-testing to be voluntary and that the testing results be confirmed by an authorized HIV test at a health facility and test kits must be properly disposed of. Tanzania’s Ministry of Health (MoH) has already started scaling up the oral HIV self-test in most regions through health facilities and community settings. According to the National AIDS Control Programme Tanzania (NASHCop), at the health facility level, HIVOST kits are provided for free at request by individuals at OPD, CTC, and TB clinics. In the context of a health facility, an individual is provided with three test kits to distribute to two other individuals to conduct HIV testing at home and bring HIV results back. At the community level, the oral HIV self-test kits are distributed by peer educators, CHWs, and healthcare workers during outreach services. The kits will also be available at selected pharmacies in the community. The HIV test results of HIVST are systematically recorded during the dispensing process of the HIVST kits, encompassing both the health facility and community settings. These HIVST results data are reported by CHWs and stored in the District Health Information Software 2 (DHIS2) database in the nearby health facilities. The DHS2 is an open-source, flexible, web-based health management information system that stores the health facilities’ information services data. The plan of the MoH, along with its bilateral and multilateral partners, is to rapidly scale up simple-to-use and low-cost oral HIV self-testing kits in urban and rural communities in the country to achieve the UNAIDS goal. To achieve one of the declared UNAIDS targets of having 95% of HIV-infected individuals aware of their status, one of the strategies that can be employed is to determine how to distribute the oral HIVST kits optimally, particularly in areas with low coverage of HIV testing services.

Mobile phones still contribute significantly to providing healthcare services[19]. It has been known that the use of text messaging as a tool to improve adherence to medication and attendance at scheduled appointments for HIV and other chronic diseases [20]. Mobile phones were used to improve community health workers’ performance in low- and middle-income countries [21]. On the other hand, studies have shown that there is limited research on the availability and use of mHealth by health workers for disease diagnosis and treatment support in sub-Saharan Africa [22]. A systematic review has shown that there are barriers to the use of mobile health in improving health outcomes in developing countries such as infrastructure, lack of equipment, and technology gap [23]. The unstructured supplementary service data (USSD)-based application can assist in the implementation of real-time tracking information [24].

The proposed program would concentrate on the door-to-door distribution of oral HIV self-test kits to male-headed households by CHWs at the community level. The male-headed households are proposed to be selected in this program because it has been shown that in Tanzania men have greater influence on decision-making processes in the context of healthcare services [25]. Furthermore, in Tanzania, male HIV testing rate is low compared to women [26]. It is known that HIV self-testing empowers users and helps normalize testing [27]. We also propose implementing a novel mobile application for self-reporting HIV test results. It is revealed that self-report provides good estimates of test-detected HIV prevalence, suggesting that it can be used when HIV test results are not available [28]. We expect quick self-reporting will translate into getting support and early initiation of care and treatment services. The program will also implement the promotion campaign of HIV self-testing and self-reporting through community engagement using the existing community structures of the CHWs platform. The program will gain experience and evidence from a pilot study conducted in a rural district in Tanzania [14]. The finding of the pilot study indicated promising results of a high acceptability rate of participants reported to have tested for HIV after the sensitization campaign. The pilot study showed a high uptake (97%) of oral HIV self-testing kits among rural study participants. Those kits were distributed by CHWs using the door-to-door approach in male-headed households [14]. Related to the HIV self-reporting results, the investigators will utilize experience obtained from a similar work published by Msuya et al 2024 on “Feasibility of a mobile application for self and assisted reporting of COVID-19 self–testing results in Tanzania: a pilot study” [29]. Several studies have been conducted in Tanzania related to HIV self-testing [30-34]. However, no studies have been conducted assessing oral HIV self-testing and HIV self-reporting results at the same time except in this study. Using the App will allow real-time data capture and transmission via messaging structures that can support contact tracing, disease surveillance, and outbreak monitoring. This program could offer digital health solutions by using the proposed App which is called *“HIV-SUSPECT APP”* for self-reporting HIV testing results among men-headed households in Tanzania. More importantly, it could enhance patient-centered care by increasing access to HIV testing. The quick test and sharing results will assist MoH in timely HIV endemic control.

We hypothesize that if CHWs use a door-to-door approach to distribute HIVOST kits and include a novel mobile application (HIV-SUSPECT APP) to report test results, then: (1) more men will test for HIV infection during program implementation, and (2) more individuals will report test results during program implementation

Therefore, the study aims to evaluate the impact of community health workers’ door-to-door distribution of oral self-testing kits and the HIV-SUSPECT APP on self-reporting of HIV testing results among men-headed households in Tanzania.

## Key definitions

<u>Acceptability of HIVST</u> is defined as the proportion of male participants who are the heads of households who will consent and accept to receive the HIVST kits for testing.

<u>Uptake of HIVST</u> is defined as the proportion of study participants who will consent and offer to test for HIV infection and report back their results.

<u>Acceptability of HIV-SUSPECT APP</u> is defined as the proportion of male participants who are the heads of households who will consent and accept the use of the HIV-SUSPECT APP for reporting.

<u>Uptake of HIV-SUSPECT APP</u> is defined as the proportion of study participants who will consent and use the HIV-SUSPECT APP for reporting back their results.

## Material and Methods

### Study design

We will use a randomized cluster study design with wards as the clusters to test a well-defined intervention using a door-to-door for the kit distribution and the HIV-SUSPECT APP. A minimum of 16 wards will be required, including 8 interventions and 8 controls. The 16 wards will be selected from the four regions: Pwani, Kagera, and Mbeya regions. These regions have low HIV testing rates as explained below in the study areas. Each region will have two selected districts and four selected wards (Figure 1). To ensure comparability of intervention and comparison wards, we will pair intervention and comparison wards based on rural and urban settings within the same district. There would be a distance from one district to another district to avoid contamination of the intervention. Thus, we will have a total of 8 pairs of intervention wards (I-ward) and control wards (C-ward) within the four regions for the study (Figure 1). After pairing the wards, we will randomize each pair so that one ward is assigned to the intervention group, and one ward is assigned to the control group.

**Figure 1:**
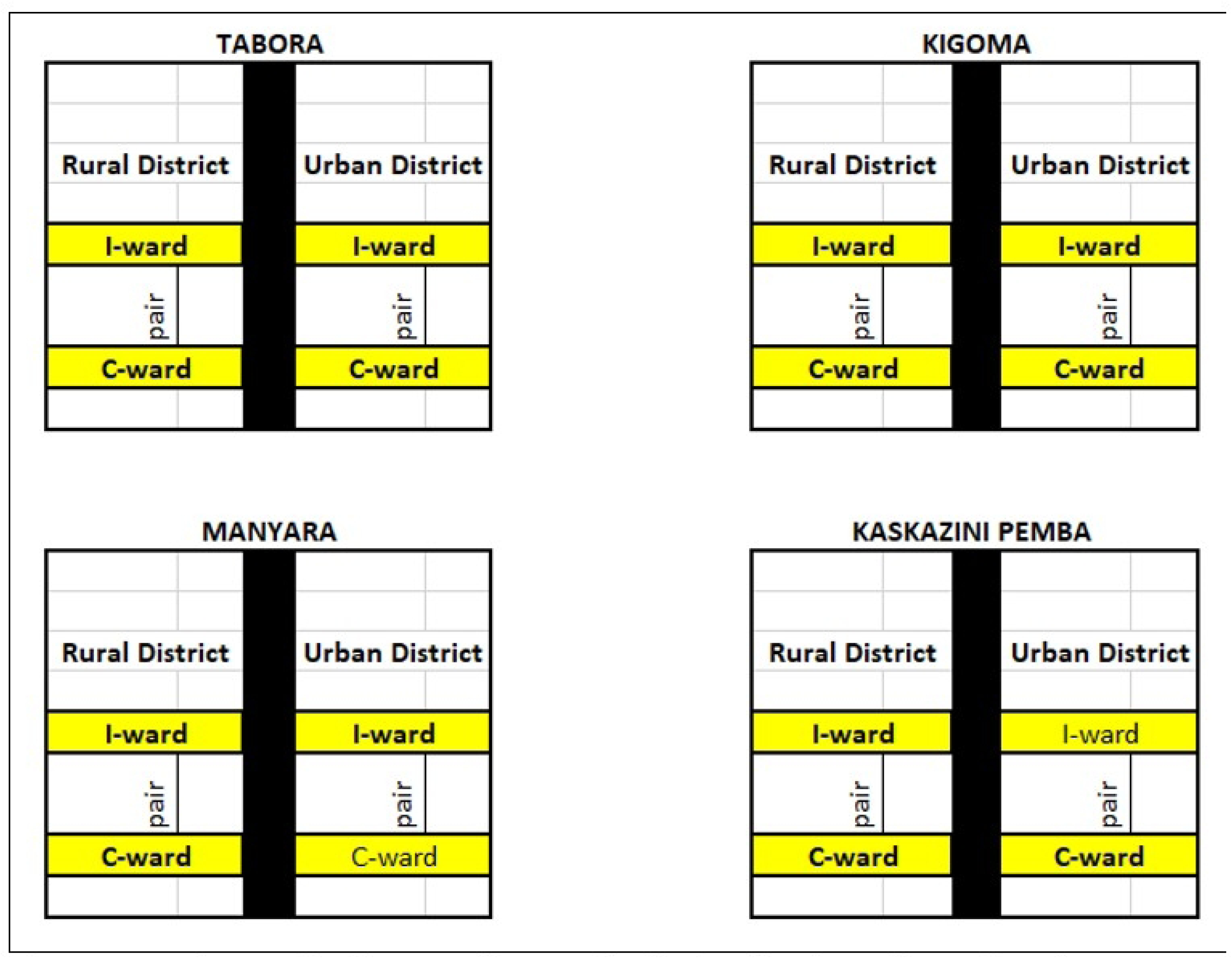
Schematic disgram for study design in the selected regions.

The baseline (pre-intervention) and follow-up survey will be conducted. At baseline, participants will complete a baseline questionnaire about demographics, HIV testing history, sexual behaviors, and mobile phone numbers. Eligible participants will receive the HIVST kits. Participants will be asked to report the results of their HIV via the HIV-SUSPECT APP. After two weeks, if the participants are unable to report their results, they will receive a non-identifying reminder text message. The follow-up survey will be conducted after two months, a brief questionnaire will be administered to assess the experience, acceptability, and uptake of the HIVST kits and the HIV-SUSPECT APP. In addition, a health facility survey will be conducted after 12 months, and data related to HIV results will be collected in the DHS2 before and after the program implementation to assess the impact of the program, will also conduct the FGDs, IDIs, and KIIs to provide qualitative information on the program to supplement survey findings.

We will invite community members (both intervention and control) to share their program experiences in focus group discussions (FGDs). These will elicit perceptions from community members on aspects of the interventions (HIVST kits and HIV-SUSPECT APP) experience and challenges faced by community members in HIV testing and HIV-SUSPECT APP.

### Promotion Campaign

A series of open meetings in the selected communities will be held as a way to engage the community strongly and to create demand for self-test/assisted testing in the participating areas. Teams of the CHWs will promote and distribute HIVOST kits self-tests to eligible members of the community. Eligibility is based on the Tanzanian law that only persons aged 18 years or above can undertake self-testing if provided with self-test kits. The promotion campaign will be as on the ongoing campaign at nationwide HIV testing campaign ***“Furaha yangu”*** literally meaning ***“My pleasure”*** meant to encourage all Tanzanians, especially men and youths to go for testing and start early treatment if found HIV positive [35]. In addition, the Information, Education, and Communication campaign will inform the respective community about the availability of HIVOST kits through the CHWs for HIV detection and a mobile App for self-reporting HIV results. mobile numbers of the CHWs will be provided/displayed in the selected communities to allow requests for HIV kits. The activities in the intervention and control wards are indicated in Table 1.

**Table 1:**
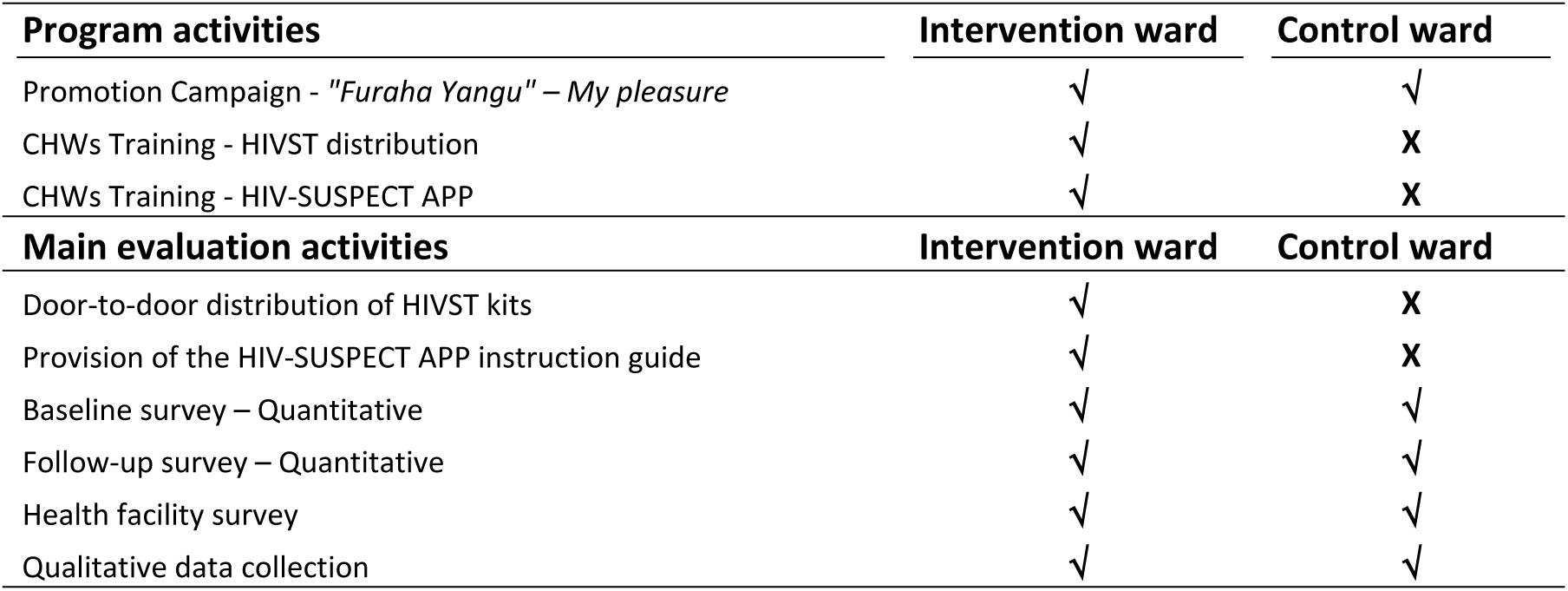
The activities in the intervention and control areas.

| **Program activities** | **Intervention ward** | **Control ward** |
| --- | --- | --- |
| Promotion Campaign - *“Furaha Yangu” – My pleasure* | √ | √ |
| CHWs Training - HIVST distribution | √ | X |
| CHWs Training - HIV-SUSPECT APP | √ | X |
| **Main evaluation activities** | **Intervention ward** | **Control ward** |
| Door-to-door distribution of HIVST kits | √ | X |
| Provision of the HIV-SUSPECT APP instruction guide | √ | X |
| Baseline survey – Quantitative | √ | √ |
| Follow-up survey – Quantitative | √ | √ |
| Health facility survey | √ | √ |
| Qualitative data collection | √ | √ |

### Study areas and population

The program will be implemented among individuals in four regions; one region from the island of Zanzibar and three from Tanzania’s Mainland that had low coverage rates of prior HIV testing among men. These regions are Tabora (50.9%), Kigoma (45%), Manyara (40.9%), and Kaskazini Pemba (31%) [26]. In these regions, partners support the delivery of comprehensive HIV services, including HIV testing, care, and prevention. Tabora - MDH, Kigoma - THPS, Manyara - EGPAF and THPS, Kaskazini Pemba – AMREF. According to the information provided on the TCRA website, it is projected that by June 2023, the regions of Tabora, Kigoma, Manyara, and Kaskazini Pemba in Tanzania will have a respective number of mobile phone users. Tabora is expected to have approximately 3,099,577 mobile phone users, Kigoma is projected to have around 1,279,021 mobile phone users, Manyara is estimated to have about 1,602,558 mobile phone users, and Kaskazini Pemba is anticipated to have approximately 99,065 mobile phone users. In addition, the percentage of men age 15–49 who own any mobile phone in Tabora (64.8%), Kigoma (65.9%), Manyara (68.1%), and Kaskazini Pemba (69.1%) [4].

In each region, the program will be conducted in four specific administrative wards. Meetings with the regional district administrative authorities will be held to select the participating wards. The activity summary of the study areas is shown in Table 2.

**Table 2:**
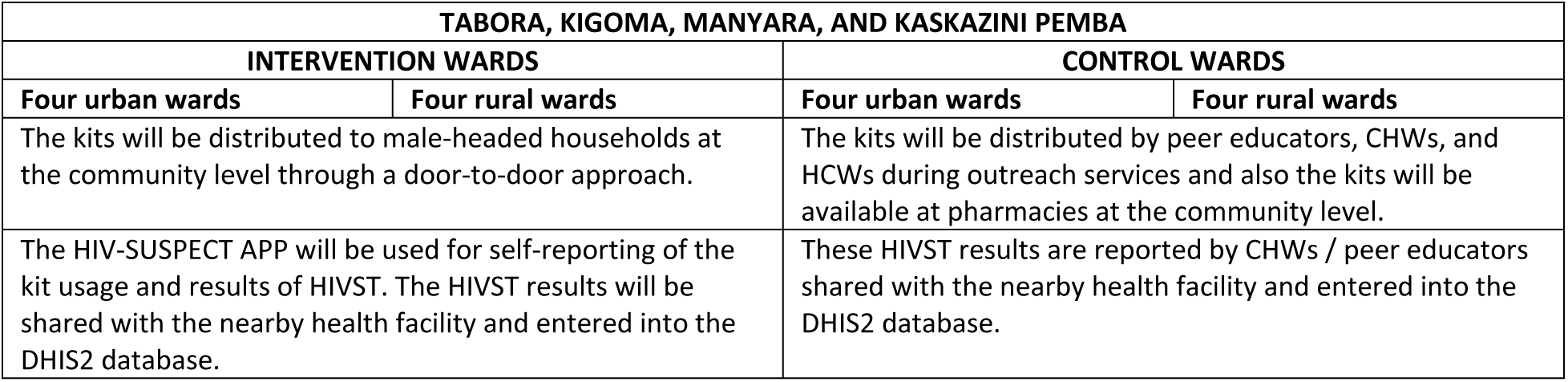
The activity summary of the study areas.

| TABORA, KIGOMA, MANYARA, AND KASKAZINI PEMBA |  |  |  |
| --- | --- | --- | --- |
| INTERVENTION WARDS |  | CONTROL WARDS |  |
| Four urban wards | Four rural wards | Four urban wards | Four rural wards |
| The kits will be distributed to male-headed households at the community level through a door-to-door approach. |  | The kits will be distributed by peer educators, CHWs, and HCWs during outreach services and also the kits will be available at pharmacies at the community level. |  |
| The HIV-SUSPECT APP will be used for self-reporting of the kit usage and results of HIVST. The HIVST results will be shared with the nearby health facility and entered into the DHIS2 database. |  | These HIVST results are reported by CHWs / peer educators shared with the nearby health facility and entered into the DHIS2 database. |  |

### Sample size calculation

The sample size calculation is based on before-after intervention with partially overlapping cohorts [36]. The operation research will be conducted in four regions Tabora, Kigoma, Manyara, and Kaskazini Pemba with population sizes of 3,391,679; 2,470,967; 1,892,502; and 272,091 respectively. The variable of interest is the coverage rate of prior HIV testing among men. The previous testing prevalence among men in Tabora, Kigoma, Manyara, and Kaskazini Pemba is 50.9%, 45%, 40.9%, and 31%, respectively [4]. The sample size (n) is equal to the following equation;

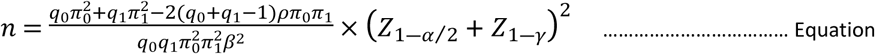

Where:

*q*_0_ and *q*_1_ are the observation probabilities

*π*_0_ and *π*_1_ are the baseline response rate the after-intervention response rate

*ρ* is the within-subject correlation

*β* is odds ratio between the after- and before-intervention responses

We assumed the intervention would result in a 50% increase in the self-testing from HIV testing coverage statistics after the program implementation in all regions. We thought the within-subject correlation of 0.15. To test the hypotheses that the intervention will result in a 50% increase in self-testing among individuals with 80% power at a 5% two-sided significance level and considering a 10% non-response rate and with a design effect of 1.5, the following sample sizes will be required, 1,210 individuals in Tabora, 1,980 individuals in Kigoma, 2,900 individuals in Manyara, and 8,780 individuals in Kaskazini Pemba. The overall sample size will be 14,870 participants in regions. The region’s estimated sample size increases with decreasing HIV testing rates. The population of the relevant selected urban and rural districts will determine how the sample size is distributed in the selected urban and rural districts. The sample size distribution in the selected regions is shown in Table 3.

**Table 3:**
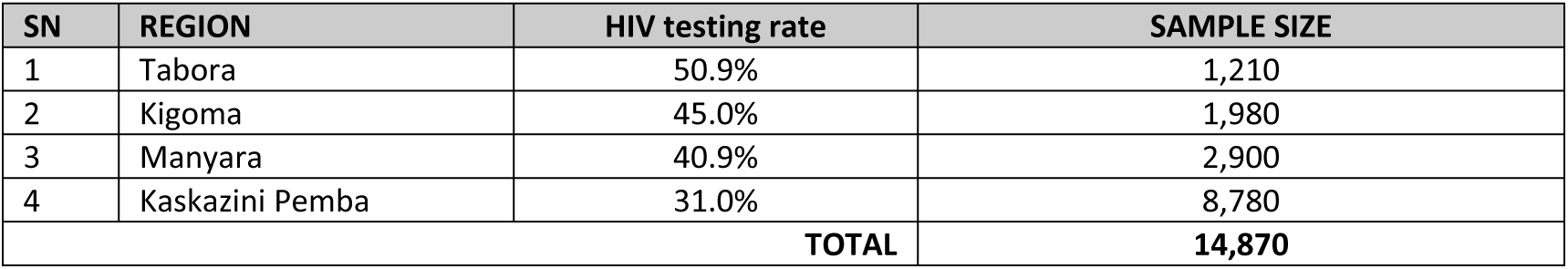
Sample size distribution in the selected regions in Tanzania.

The sample size for qualitative studies will be determined by reaching thematic saturation, that is, adherence to the principle of saturation and consideration of the evaluation needs [37]. The qualitative sample size aims to achieve data saturation, where no additional emerging or re-emerging themes are identified. However, if data saturation is not reached within a specific group, additional interviews will be conducted until saturation is achieved. In qualitative assessment, we will conduct a total of 2-4 FGDs at each of the districts including male and female participants. Each FGD will include approximately 8 participants. In addition, 3-4 In-depth Interviews (IDIs) in each district and 27-28 Key Informant Interviews (KII) will be done. The sample size distribution for the qualitative approach is shown in Table 4.

**Table 4:** Summary of qualitative sample size.

| Sample type | Method of data collection | Total |
| --- | --- | --- |
| **Key officials** |  |  |
| Ministry of Health – NASHCop staff | KII | 3 |
| Regional AIDS Coordinator (RAC) | KII | 4 |
| District AIDS Coordinator (DAC) | KII | 8 |
| Regional Medical Officer (RMO) | KII | 4 |
| District Medical Officer (DMO) | KII | 8 |
| Stakeholders in the relevant regions | KII | 4 |
| Health care providers | IDIs | 24 |
| **Community members** |  |  |
| FGD in the Intervention districts (Males and Females) | FGD | 8 |
| FGD in the Control districts (Males and Females) | FGD | 8 |

### Sampling procedure

In quantitative, for the three regions from Tanzania Mainland, a four-stage stratified cluster sampling technique will be used to select participants. In the first step, two districts—one rural and one urban— will be chosen at random from each region. The second stage will involve randomly selecting one ward in each district. The third stage will involve the selection of the enumeration areas (EAs). The National Bureau of Statistics (NBS) will be asked to provide the list of enumeration areas and respective wards. The C-Survey 2.0 software will be used to perform the EAs selection using the probability proportional to size (PPS) method. The fourth stage will systematically select male-headed households with eligible study participants in each EA. In one region from Zanzibar, a three-stage stratified cluster sampling technique will be used to select participants. Deliberately, only two districts in Kaskazini Pemba will be included. The first stage will involve randomly selecting one shehia in each district. The second stage will involve the selection of the EAs. The third stage will systematically select male-headed households with eligible study participants in each EA. At least two household members will be required to participate in the survey, Figure 2.

**Figure 2:**
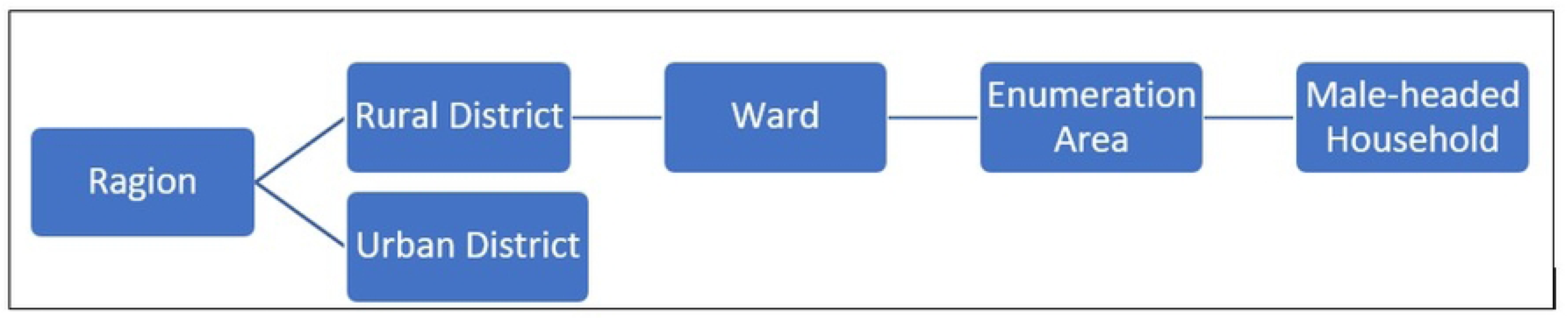
Sampling approach.

In qualitative, the community members for FGD sessions will be selected purposely, especially those with experience in using interventions in their households. The IDIs will be conducted among healthcare providers in selected health facilities in each district (district hospital, health center, and dispensary). The KIIs will be conducted among Regional Medical Officers (RMOs), District Medical Officers (DMOs), NASHCop staff, Regional AIDS Coordinators (RACs), District AIDS Coordinators (DACs), and other stakeholders.

### HIVST algorithm

According to the WHO [38], all reactive HIVST results will be followed by further testing by a trained healthcare provider to confirm HIV status. Non-reactive HIVST results will be considered HIV-negative, with no need for immediate further testing. Those with unknown HIVST results need to repeat the test using another HIVST kit or seek testing from a CHW. Any person uncertain or unknown about their HIVST result will be encouraged to seek testing from a trained healthcare provider, Figure 3.

**Figure 3:**
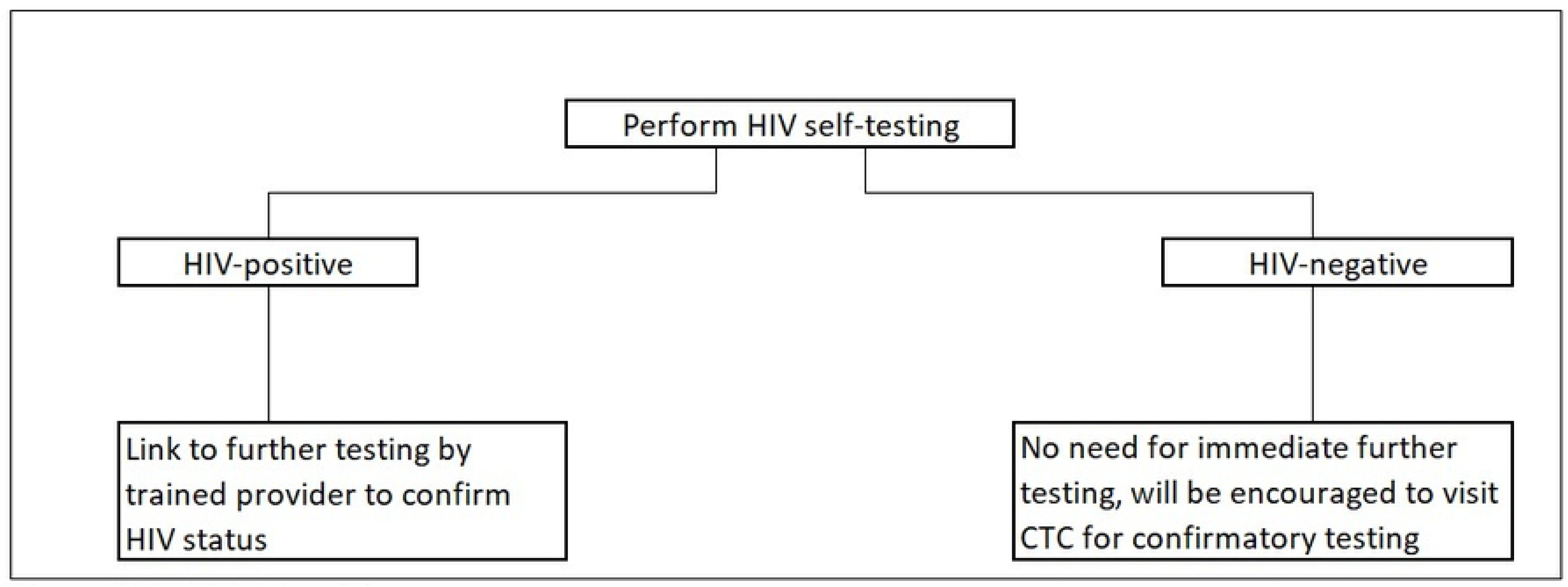
HIVST algorithm.

### Study outcomes

Primary outcomes;

1. The proportion of men who tested for HIV infection by oral HIV self-testing.
2. The proportion of individuals who reported HIV test results using an HIV-SUSPECT APP.
3. The proportion of newly diagnosed HIV-positive individuals who will link into CTC.

Secondary outcomes;

1. The proportion of all individuals tested for HIV during program implementation by any testing method.
2. The proportion of all individuals reported HIV test results before and after the program implementation.

### Data collection

The data collection will take place for 9 months, Table 5. All adult individuals aged 18 years old or above from selected wards will be eligible for inclusion in the program. In addition to age, other inclusion criteria will be individuals with the following characteristics; never tested for HIV or tested HIV three months ago or more, HIV status: negative, don’t know/don’t remember or test results were unknown, live in selected ward, and provide informed written consent to participate in the study. The individuals must be from male-headed households with at least one mobile phone.

**Table 5:**
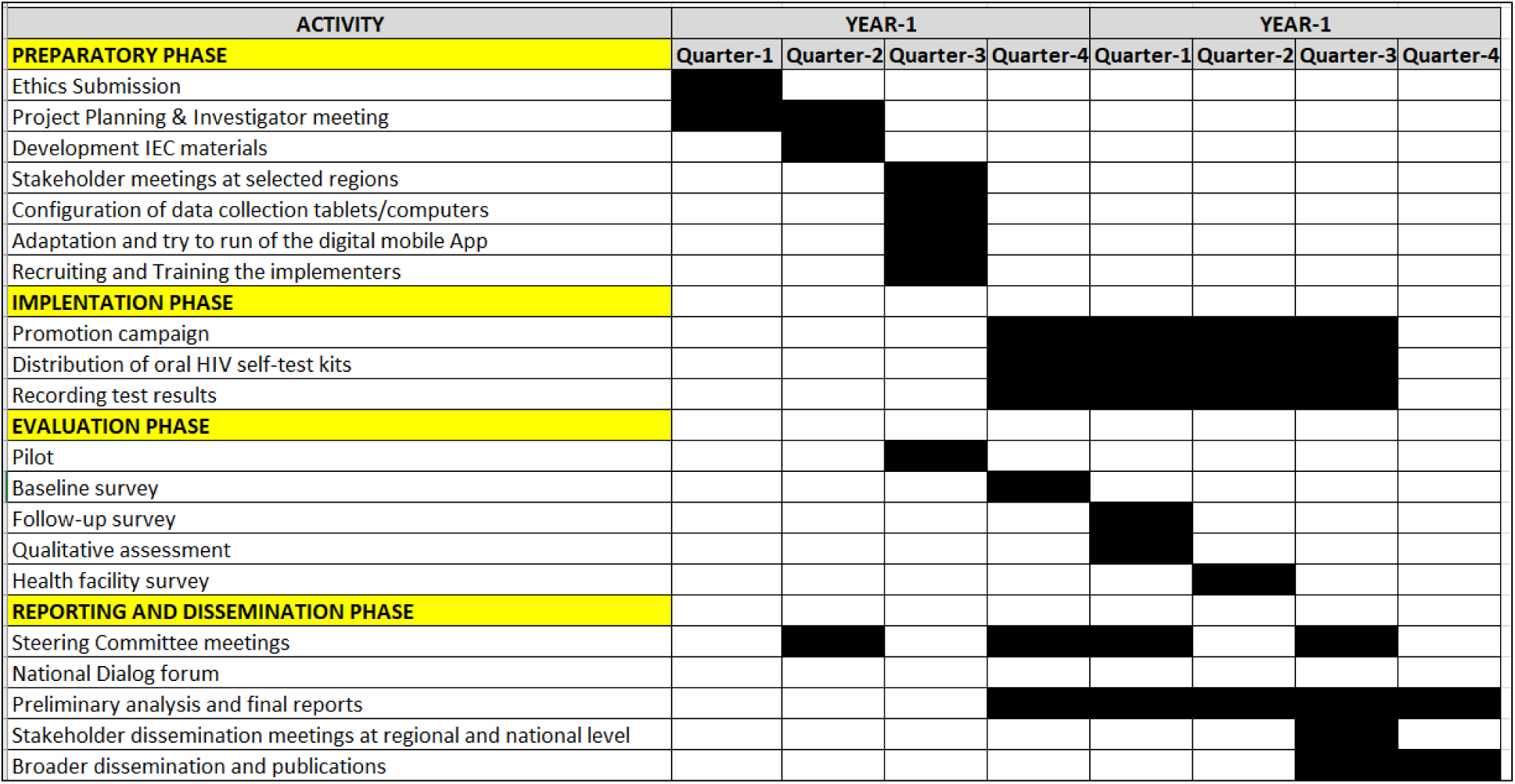
The details of the activity plan.

The program evaluation will use quantitative and qualitative data to assess program implementation. A research assistant (RA) will be positioned in the selected wards to oversee the evaluation activities. Prospective surveys will be embedded in the participating wards at the beginning, midway, and end of the program in intervention and the control wards in all selected regions. Before actual intervention, the pilot program will be carried out to fine-tune the intervention activities before its full implementation. The pilot program wards will be different from the intervention wards in each region. CHWs will provide a demonstration of how to use HIVOST kits using the instructional video. They will conduct a live demonstration of how to use and interpret results. Community members will be provided with a choice to take a kit home or to use it onsite either on their own or with the assistance of peer educators, CHWs, or healthcare workers (HCWs). Members of the community who choose to test on-site will be able to verify reaction results at the local health facility. During the pilot program, oral consent will be sought from those who agree to do the HIV test themselves and provide a mobile number to record the person receiving the test and for the MoH to enquire about the results for appropriate action and do a quality check of the program.

#### a) Quantitative assessments

A research assistant (RA) will work closely with the CHWs of the specific selected ward. The eligible individuals at the household level will be given study information and asked for written consent to participate in the evaluation. The baseline questionnaire will assess barriers and facilitators to HIV testing, gender power imbalance dynamics within the household, HIV testing history, HIV knowledge, and basic sociodemographic information including education, employment, medical history, and measures of household wealth. The six parameters will be included in the HIV-SUSPECT APP; age in categories: (18-25) (25-39) (40-59) (60˃), sex: (Female/Male), Identity: (Phone number/3 initials), location: (Regional/District/ Ward/House number), who performs the test: (Self-Test/household member/CHW), and test results in HIV status: (Positive / Negative / Indeterminate).

The follow-up survey will assess testing acceptability, barriers, and facilitators to testing, testing results, disclosure of results, social outcomes of testing, linkage to care among those testing positive, and details of the HIVOST kits use. The implementation expenditures outlined by inputs will be tracked and combined with assessment data to estimate the intervention cost of the door-to-door approach and the mobile application. All data will be collected using semi-structured questionnaires loaded on tablets except for the consent forms will be on paper. Participants will be asked to report the results via a mobile App or directly assisted by RA or CHW in conducting the HIVST report.

#### b) Economic cost assessments

The team will collect and analyse primary data from the study sites using standardized questionnaires. The program expenditures, supplemented by on-site observation and assessments evaluation data will be used to estimate total economic and unit costs of HIVOST kits distribution using the door-to-door approach and a novel mobile application, by input and site (region). Inputs will be categorized into start-up, capital, and recurrent costs. Sensitivity and scenario analyses will be performed to assess the impact of key parameters on unit costs.

#### c) Qualitative assessments

The implementation and expansion of HIVST must be carefully tailored to the local context to maximize its utility. We will explore the cognitive and socio-cultural adaptation of the use of HIVOST kits in rural and urban community settings in Tanzania. The qualitative study will be used to capture understanding and knowledge of HIVOST kits as well as assess issues that will promote access to and use of HIVOST kits in the communities through FGDs, IDIs, and KIIs. The FGDs will be conducted with the community members. This method serves to solicit general attitudes and perceptions, knowledge, experiences, and practices, shared in the course of interaction among the community. Furthermore, the KIIs will be conducted to gather their perspectives and understanding as well as the socio-cultural ideas on the use and adaptation of the HIVOST kits deployment in Tanzania. The IDIs will be conducted to get a deeper insight understanding of the selected topics of the HIVOST kits, experiences of harnessing existing platforms to accommodate HIVST, factors affecting the uptake of HIVOST kits, constraints, and enablers as well as social-cultural factors that promote the uptake of HIV oral self-tests.

### HIV-SUSPECT APP-USSD Operation

The HIV-SUSPECT APP-USSD will be hosted on the *<u>Beem USSD Hub</u>* which provides an aggregated connection to all mobile network operators and an application programming interface (API) to manage the menu and web dashboard for displaying the data collected from the USSD responses. The *<u>Beem USSD Hub</u>* offers a single unified API to manage and run a USSD service across multiple mobile networks from one point. It functions as a broker among the subscriber, mobile operator, and customer. When a USSD session is initiated, the hub calls the HIV-SUSPECT APP-USSD to retrieve the response and return it to the subscriber or study participants. This process continues until the session times out or is terminated either by the HIV-SUSPECT APP-USSD or by the subscriber or study participants themselves. A session must be allocated for every transaction request; the response for this request and the subsequent series of requests and responses within that session all share the same session ID until the session is closed or times out, as reported elsewhere regarding USSD operations [39]. The security will be offered by the HIV-SUSPECT APP-USSD to study participants. Compared with SMS, the HIV-SUSPECT APP-USSD is considered relatively more secure because no copy of the message is stored on the study participant’s mobile phone. A single session is established between the mobile terminal and the application server, and at the USSD gateway, the message is encrypted, preventing data from being misused between the gateway and the server.

### Link to care and treatment clinic

Individuals will be offered HIV oral self-testing kits with detailed pre-test counseling for use of the kit and actions to take in response to a positive or negative test or unknown. The pre-test counseling will include basic HIV education, benefits of testing, and disclosure. Participants will be advised to seek confirmatory testing at the care and treatment clinic (CTC) to undergo the confirmatory HIV tests if the self-test result is positive or unknown results. Those who tested positive will be treated based on standard procedures of the CTC. Individuals who tested negative will be also encouraged to visit CTC for confirmatory testing. The program team will make close follow-up to those who reported positive by oral HIV self-testing via their mobile phones. The program team will call back to those individuals via their shared mobile numbers. The program team will ask them if those individuals visited the CTC. The program staff will inquire about information visiting the CTC. The inquiry information will be: Did you request confirmatory testing at a nearby CTC? If, yes, what is your test result? If, positive, did you visit a nearby CTC for HIV services? Were you initiated on antiretroviral therapy (ART)? If the individual is positive and did not attend CTC for HIV services, she/he will be visited by CHW to encourage the individual to seek HIV services as soon as possible.

### Data management and statistical analysis

#### Quantitative data

Tablet-based open data kit (ODK) forms will be used to capture study information and managed via the electronic database of the REDCap platform. Data will be extracted from questionnaires and will be entered and managed in an electronic database. All data will be collected, stored, and maintained within IHI saver in Dar es Salaam Tanzania. Statistical analyses will mainly involve descriptive analysis of the primary outcome of acceptability and uptake of the interventions. Comparison will be made between before and after the intervention. Descriptive summaries will be prepared for tests offered, tests refused, self-tests done, assisted testing done, and test results based on persons offered tests in communities. Internal comparison between intervention ad control areas for these parameters. Logistic regression modelling will be done for the analysis of the predictors of oral HIV self-test uptake as well as the performance of the selection criteria. Secondary analysis of the proportion of tests conducted and reported in the HIV -SUSPECT app will also be done and other variables.

The costing analysis will use the provider perspective to estimate resources used to deliver HIVST intervention [40-43]. The ingredients-based costing mainly at the service delivery and intermediate administrative level covering the cost of all key program activities such as training, social mobilization, transport, storage, and waste management from the implementation and control area will be captured. This will be done hand in hand with mapping of the service delivery process, key funding flows and funding sources relevant to the different HIVST delivery activities. Also captures the source and use of various types of human resources for health (HRH) to deliver HIVST, including the CHW, peer educators, and HCWs [44].

#### Qualitative data

The FGDs, IDIs, and KIIs will be recorded using an MP3 voice recorder upon participants’ consent. After the FGD, the note-taker and the moderator will review their hand-written notes for quick feedback. After revisions, the notes will be typed, saved, and exported to the Nvivo program for analysis. Qualitative data will be transcribed verbatim. Quality checks will be done by listening to audio interviews versus the transcripts to identify and correct missing and typing errors respectively. The transcripts will be exported into NVIVO analysis software to analyze data focusing on emerging themes, patterns, similarities, and differences. We will use open coding to label concepts and define and develop categories based on the properties and dimensions of participants’ descriptions. We will use both inductive (ideas emanating from the data itself) and deductive (theoretical understanding, literature review, and researcher’s experience) approaches for data analysis. Analyses will be performed using qualitative data analysis software - NVIVO version 12.

## Ethics and dissemination

This protocol has been reviewed and approved by the Human Research Ethics Committee of the Ifakara Health Institute - Institutional Review Board (IHI-IRB) with reference number: IHI/IRB/No: 14-2025. During recruitment, we will obtain written informed consent from participants. We will provide potential participants with appropriate information about the study and their involvement in the study so they can make an informed choice. Potential participants in the households will be briefed about the nature and goal of the study, as well as the anticipated risks and advantages, at the beginning of each interview. All potential participants will be made aware that their participation is voluntary and does not affect their eligibility to receive services from any health services now or in the future. All participants, both literate and illiterate will be informed that the data collected will be held in strict confidence. To ensure that participants are aware that the survey includes questions on highly personal and sensitive topics, the interviewer will warn them that some topics are difficult to talk about. The respondent will be made aware at the outset that he or she is free to terminate the interview at any point and to skip any questions that he or she does not wish to respond to. Participants will be offered to keep a copy of the written consent form.

Upon completion of data collection, the Scientific Steering Committee members will participate in a data interpretation workshop to review preliminary results and discuss implications. The feedback from this interpretation workshop will then inform the final report, and a national-level dissemination meeting will be held at the end of the project to inform Tanzania stakeholders of the findings. Furthermore, if funding is available, we will collaborate with MoH representatives through the National Aids Control Program (NASHCop). So, we’ll plan and incorporate findings obtained into the relevant policy in the health system for the sustainability of the program. The findings will be also disseminated through appropriate media, conferences, meetings, online forums, and publications.

## Quality assurance and quality control

The study will be conducted by procedures identified in the protocol and relevant Standard Operation Procedures (SOPs) which will specify the types of materials to be reviewed, the responsible party, and the schedule for reviews. Study-specific training will be provided for all staff before the commencement of the program. Monitoring of the quality of operations and integrity of data collected will be performed by the quality assurance team from IHI. Hence, following written SOPs, the quality assurance team will verify that the program is conducted and data are generated, documented (recorded), and reported in compliance with the protocol, GCP, and the applicable regulatory requirements. Investigators will provide direct access to source data/documents, and reports for monitoring and auditing by the quality assurance team and inspection by local ethics and regulatory authorities.

## Activities timeline

This program will be completed in 24 months Table 5 below shows the details of the activity plan.

## Financial Considerations

The authors received no specific funding for this work. The authors are looking for interested funders to finance the program.

## Discussion

The program intervention will involve CHWs providing at least three HIV Oral Self-Test (HIVOST) kits to male-headed families in the selected households, that is, *the door-to-door distribution of oral self-testing kits by Community Health Workers.* Within specific selected wards, trained CHWs will move from one male-headed household to another and offer HIVOST kits to eligible clients. Individuals will be provided with the HIVOST kits with the option of either being directly assisted (in the presence of a CHW) or unassisted using an instructional video that will be given to them. The CHWs will share their mobile numbers with the households’ heads. This will help to get support whenever needed such as more kits. The program will work with the Health Management Teams in the targeted areas to prepare an implementation plan with the program team. Training of CHWs will be done on the distribution of the HIVOST kits and performing testing using the instructional video ([video-link](https://www.youtube.com/watch?v=gfsmnMVC8Ug)). The video developed by MoH on the procedure of the HIVOST. The MoH will manage the implementation of the training to ensure that the process will be sustainable and the rollout is smooth beyond the program implementation period. A similar distribution model of the door-to-door distribution of oral self-testing kits was used in rural Lesotho [45], Zambia [46], Malawi [47], and South Africa [48].

During the program implementation, the HIV-SUSPECT App will be adopted for self-reporting of HIV test results. Individuals participating in testing activities can enter their mobile numbers, area codes, and test results into the system. Those individuals given the test kits will be requested to fill in the test results in the App or inform the CHW to do that. The individuals in the same household both males and females can enter the test results using one mobile phone. As pointed out above, this application was utilized in the earlier COVID-19 antigen quick diagnostic self-testing research pilot deployment, where participants assisted or self-reported their test results [29]. After the kits are distributed, an automated reminder message will be sent out within 24 hours encouraging people to get tested for HIV, utilize the App, and self-report their results. The mobile App instruction guide, the self-pre-test counseling guide for HIV testing, an instructional video, and HIVST kits with written instructions are all part of the intervention package that will be distributed to all visited male-headed households. Mobile Apps were also used in various countries. In Ghana, for example, people were able to self-report their COVID-19 symptoms and whereabouts through an online platform that was enabled by the USSD protocol [49]. In Tanzania, mobile phone-based solutions have been employed to solve the difficulties of monitoring and supervising a large number of geographically dispersed health workers to increase the reach and effectiveness of field health workers [50].

The program will be evaluated by cluster randomized design. A key strength of cluster randomized designs is their ability to evaluate interventions that are naturally delivered at the group level, like community programs or policy changes, while also minimizing contamination between treatment and control groups [51].

Despite the strengths of the cluster randomized design, there are a few limitations. First, the cluster randomized design is useful for evaluating the proposed interventions at the group level, but it has limitations including requiring larger sample sizes and potentially higher costs compared to individual randomization, and increased complexity in design, analysis, and implementation [52]. Second, this study excludes potential participants who do not have mobile phones.

In conclusion, this program will offer useful resources for other researchers to implement comparable procedures in different contexts for HIVOST kit distribution and HIV result self-reporting. The lack of funding has prevented the implementation of this protocol. Therefore, any interested sponsor or funder is invited to work with us to conduct this study in Tanzania.

## Data Availability

No datasets were generated or analysed during the current study. All relevant data from this study will be made available upon study completion.

## Acknowledgments

The protocol was critically reviewed by the Ifakara Health Institute Review Board, for which the authors are grateful.

## Author Contributions

**Conceptualization:** Abdallah Mkopi

**Data curation:** Silas Temu.

**Formal analysis:** Ali Ali.

**Methodology:** Hajirani Msuya, Kassimu Tani, Omar Lweno, Abdallah Mkopi

**Writing – original draft:** Hajirani Msuya and Abdallah Mkopi.

**Writing – review & editing:** Kassimu Tani, Omar Lweno, Mwifadhi Mrisho, Bakar Fakih, Omar Juma, Ali Ali, Silas Temu, Gumi Abdallah, Hassan Tearish, Zeye Masunga, Werner Maokola, Neema Mpanduji, Moza Kassim, Ramadhani Abdul

## Notes

### Competing Interest Statement

The authors have declared no competing interest.

### Clinical Trial

"N/A"

### Funding Statement

The author(s) received no specific funding for this work.

### Author Declarations

On 28th March 2025, the Ifakara Health Institute Review Board (IHI-IRB) reviewed from study titled: "Study protocol for evaluation of the impact of the door-to-door approach by community health workers and the novel mobile application for self-reporting on HIV testing rates among men-headed households in Tanzania", submitted by the Principal Investigator, Dr Abdallah Mkopi. The study has been approved for implementation after IRB consensus

## Reference

1. Amuche NJ, Emmanuel EI, Innocent NE. HIV/AIDS in sub-Saharan Africa: current status, challenges and prospects. 2017.

2. Frescura L, Godfrey-Faussett P, Feizzadeh A A, El-Sadr W, Syarif O, Ghys PD, et al. Achieving the 95 95 95 targets for all: A pathway to ending AIDS. PLoS One. 2022;17(8):e0272405.

3. Chamie G, Napierala S, Agot K, Thirumurthy H. HIV testing approaches to reach the first UNAIDS 95% target in sub-Saharan Africa. The lancet HIV. 2021;8(4):e225–e36.

4. MoH N, OCGS, and ICF. Tanzania Demographic and Health Survey and Malaria Indicator Survey 2022 Final Report. Dodoma, Tanzania and Rockville, Maryland, USA: Ministry of Health (MoH) [Tanzania Mainland], Ministry of Health (MoH) [Zanzibar], National Bureau of Statistics (NBS), Office of the Chief Government Statistician (OCGS), and ICF, 2022.

5. Njau BJ. A multi-component theory-based behaviour change intervention to increase HIV self–testing uptake and linkage to HIV prevention, care and treatment among hard to reach adults in Northern Tanzania. 2021.

6. Pai N, P, Dheda K. HIV self-testing strategy: the middle road. Expert Rev Mol Diagn. 2013;13(7):639–42. Epub 2013/09/26. doi: 10.1586/14737159.2013.820543. PubMed PMID: 24063389.

7. Njau B, Covin C, Lisasi E, Damian D, Mushi D, Boulle A, et al. A systematic review of qualitative evidence on factors enabling and deterring uptake of HIV self-testing in Africa. BMC public health. 2019;19:1–16.

8. Olakunde BO, Alemu D, Conserve DF, Mathai M, Mak’anyengo MO, Group NS, et al. Awareness of and willingness to use oral HIV self-test kits among Kenyan young adults living in informal urban settlements: a cross-sectional survey. AIDS care. 2023;35(9):1259–69.

9. Bbuye M, Muttamba W, Nassaka L, Nakyomu D, Taasi G, Kiguli S, et al. Factors associated with linkage to HIV care among oral self-tested HIV positive adults in Uganda. HIV/AIDS-Research and Palliative Care. 2022:61–72.

10. Shaba F, Balakasi K, Offorjebe OA, Nyirenda M, Wong VJ, Gupta SK, et al. Facility HIV self-testing in outpatient departments: An assessment of characteristics and concerns of outpatients who opt-out of testing in Malawi. JAIDS Journal of Acquired Immune Deficiency Syndromes. 2022:10.1097.

11. Mwansa E. Factors influencing the use of HIV self-testing kits among adolescents: a case of Kalingalinga compound, Lusaka: The University of Zambia; 2024.

12. TSHUMA MO. AN EXPLORATION OF THE PERCEPTIONS AND EXPERIENCES OF COMMUNITY MEMBERS ON ACCEPTABILITY AND FEASIBILITY OF HIV SELF-TESTING ORAL FLUID TEST IN NETA WARD MBERENGWA DISTRICT, ZIMBABWE: School of Community and Health Sciences, University of the Western Cape; 2018.

13. Choko A, T, Desmond N, Webb E, L, Chavula K, et al. The uptake and accuracy of oral kits for HIV self-testing in high HIV prevalence setting: a cross-sectional feasibility study in Blantyre, Malawi. PLoS Med. 2011;8(10):e1001102. Epub 2011/10/13. doi: 10.1371/journal.pmed.1001102. PubMed PMID: 21990966; PubMed Central PMCID: PMCPMC3186813.

14. Mkopi A, Korte J, E, Lesslie V, diNapoli M, Mutiso F, et al. Acceptability and uptake of oral HIV self-testing among rural community members in Tanzania: a pilot study. AIDS Care. 2023;35(9):1338–45. Epub 2023/05/28. doi: 10.1080/09540121.2023.2217376. PubMed PMID: 37245239.

15. Afzal MM, Pariyo GW, Lassi ZS, Perry HB. Community health workers at the dawn of a new era: 2. Planning, coordination, and partnerships. Health Research Policy and Systems. 2021;19(3):1–17.

16. Lehmann U, Sanders D. Community health workers: what do we know about them. The state of the evidence on programmes, activities, costs and impact on health outcomes of using community health workers Geneva: World Health Organization. 2007;42.

17. Mwai GW, Mburu G, Torpey K, Frost P, Ford N, Seeley J. Role and outcomes of community health workers in HIV care in sub-Saharan Africa: a systematic review. Journal of the International AIDS Society. 2013;16(1):18586.

18. NACP. NATIONAL COMPREHENSIVE GUIDELINES ON HIV TESTING SERVICES. Dodoma, Tanzania: Ministry of Health, Community Development Gender Elderly and Children National AIDS Control Programme (NACP), 2021.

19. Kannisto KA, Koivunen MH, Välimäki MA. Use of mobile phone text message reminders in health care services: a narrative literature review. Journal of medical Internet research. 2014;16(10):e222.

20. Mbuagbaw L, Mursleen S, Lytvyn L, Smieja M, Dolovich L, Thabane L. Mobile phone text messaging interventions for HIV and other chronic diseases: an overview of systematic reviews and framework for evidence transfer. BMC health services research. 2015;15:1–16.

21. Feroz A, Jabeen R, Saleem S. Using mobile phones to improve community health workers performance in low-and-middle-income countries. BMC public health. 2020;20:1–6.

22. Osei E, Kuupiel D, Vezi PN, Mashamba-Thompson TP. Mapping evidence of mobile health technologies for disease diagnosis and treatment support by health workers in sub-Saharan Africa: a scoping review. BMC Medical Informatics and Decision Making. 2021;21:1–18.

23. Kruse C, Betancourt J, Ortiz S, Luna SMV, Bamrah IK, Segovia N. Barriers to the use of mobile health in improving health outcomes in developing countries: systematic review. Journal of medical Internet research. 2019;21(10):e13263.

24. Dabas A, Dabas C. Implementation of Real Time Tracking using Unstructured Supplementary Service Data. World Academy of Science, Engineering and Technology. 2009;54:p241.

25. Shayo EH, Norheim OF, Mboera LE, Byskov J, Maluka S, Kamuzora P, et al. Challenges to fair decision-making processes in the context of health care services: a qualitative assessment from Tanzania. International journal for equity in health. 2012;11:1–12.

26. Demographic T. health survey and malaria indicator survey (TDHS-MIS) 2021–22. 2022.

27. Wood BR, Ballenger C, Stekler JD. Arguments for and against HIV self-testing. HIV/AIDS-Research and Palliative Care. 2014:117–26.

28. Chasimpha SJ, Mclean EM, Dube A, McCormack V, dos-Santos-Silva I, Glynn JR. Assessing the validity of and factors that influence accurate self-reporting of HIV status after testing: a population-based study. AIDS (London, England). 2020;34(6):931–41.

29. Msuya HM, Ali AM, Mrisho M, Lweno ON, Temu SG, Msuya I, et al. Feasibility of a Mobile Application for Self and Assisted Reporting of Coronavirus Disease 2019 Self-Testing Results in Tanzania: A Pilot Study. The American journal of tropical medicine and hygiene. 2025:tpmd240161.

30. Conserve DF, Bay C, Kilonzo MN, Makyao NE, Kajula L, Maman S. Sexual and social network correlates of willingness to self-test for HIV among ever-tested and never-tested men: implications for the Tanzania STEP project. AIDS care. 2019;31(2):169–76.

31. Nnko S, Nyato D, Kuringe E, Casalini C, Shao A, Komba A, et al. Female sex workers perspectives and concerns regarding HIV self-testing: an exploratory study in Tanzania. BMC Public Health. 2020;20:1–9.

32. Conserve DF, Muessig KE, Maboko LL, Shirima S, Kilonzo MN, Maman S, et al. Mate Yako Afya Yako: formative research to develop the Tanzania HIV self-testing education and promotion (Tanzania STEP) project for men. PloS one. 2018;13(8):e0202521.

33. Aloni MS. Drivers of HIV self-test kit among Tanzanian men aged 15–49: findings from the 2022 TDHS-MIS cross-sectional study. AIDS Research and Therapy. 2025;22(1):3.

34. Matovu JK, Mbita G, Hamilton A, Mhando F, Sims WM, Thompson N, et al. Men’s comfort in distributing or receiving HIV self-test kits from close male social network members in Dar Es Salaam, Tanzania: baseline results from the STEP project. BMC Public Health. 2021;21:1-11.

35. Conserve DF, Msofe J, Issango J, Tureski K, McCarthy P, Rwezahura P, et al. Development, implementation, and scale up of the national furaha yangu campaign to promote HIV test and treat services uptake among men in tanzania. American Journal of Men’s Health. 2022;16(2):15579883221087838.

36. Zhang S, Cao J, Ahn C. Sample size calculation for before–after experiments with partially overlapping cohorts. Contemporary clinical trials. 2018;64:274–80.

37. Vasileiou K, Barnett J, Thorpe S, Young T. Characterising and justifying sample size sufficiency in interview-based studies: systematic analysis of qualitative health research over a 15-year period. BMC medical research methodology. 2018;18:1–18.

38. WHO. Brief Policy, WHO Recommends HIV Self-Testing–Evidence Update and Considerations for Success; 2019. Geneva, Switzerland: World Health Organization, 2019 Contract No.: WHO/CDS/HIV/19.36.

39. Krugel GT. Mobile Banking Technology Options. Finmark trust. 2007.

40. McBain RK, Jordan M, Kapologwe NA, Kagaayi J, Kiracho EE, Nandakumar AK. Costing of HIV services, uganda and united republic of tanzania. Bulletin of the World Health Organization. 2023;101(10):626.

41. Organization WH. Costing guidelines for HIV prevention strategies. Costing guidelines for HIV prevention strategies 2000. p. 136-.

42. Ahmed N, Ong JJ, McGee K, d’Elbée M, Johnson C, Cambiano V, et al. Costs of HIV testing services in sub-Saharan Africa: a systematic literature review. BMC infectious diseases. 2022;22(Suppl 1):980.

43. Mangenah C, Mwenge L, Sande L, Ahmed N, d’Elbée M, Chiwawa P, et al. Economic cost analysis of door-to-door community-based distribution of HIV self-test kits in Malawi, Zambia and Zimbabwe. Journal of the International AIDS Society. 2019;22:e25255.

44. Sande LA, Matsimela K, Mwenge L, Mangenah C, Choko AT, d’Elbée M, et al. Costs of integrating HIV self-testing in public health facilities in Malawi, South Africa, Zambia and Zimbabwe. BMJ global health. 2021;6(Suppl 4):e005191.

45. Amstutz A, Lejone TI, Khesa L, Muhairwe J, Bresser M, Vanobberghen F, et al. Home-based oral self-testing for absent and declining individuals during a door-to-door HIV testing campaign in rural Lesotho (HOSENG): a cluster-randomised trial. The lancet HIV. 2020;7(11):e752–e61.

46. Mulubwa C, Hensen B, Phiri MM, Shanaube K, Schaap AJ, Floyd S, et al. Community based distribution of oral HIV self-testing kits in Zambia: a cluster-randomised trial nested in four HPTN 071 (PopART) intervention communities. The lancet HIV. 2019;6(2):e81–e92.

47. Indravudh PP, Fielding K, Chilongosi R, Nzawa R, Neuman M, Kumwenda MK, et al. Effect of door-to-door distribution of HIV self-testing kits on HIV testing and antiretroviral therapy initiation: a cluster randomised trial in Malawi. BMJ global health. 2021;6(Suppl 4):e004269.

48. Majam M. Progress on the scaling up of HIV testing in South Africa through varied distribution models using the oral HIV self-test kit. Oral diseases. 2020;26:137–40.

49. Oyejide AJ, Akinlabi A, Atoyebi EO, Falola PB, Awonusi AA, Owolabi F. COVID-19 crisis era; Engineering interventions in Sub-Saharan Africa. Nigerian Journal of Technology. 2023;42(3):389–98.

50. DeRenzi B, Borriello G, Jackson J, Kumar VS, Parikh TS, Virk P, et al. Mobile phone tools for field-based health care workers in low-income countries. Mount Sinai Journal of Medicine: A Journal of Translational and Personalized Medicine. 2011;78(3):406–18.

51. Farrugia P. Statistics in Brief: The Cluster Randomized Controlled Trial—What Is It and Why Is It Relevant to Research in Surgery? Clinical Orthopaedics and Related Research®. 2021;479(8):1852–7.

52. Hayes RJ, Moulton LH. Cluster randomised trials: Chapman and Hall/CRC; 2017.

